# Feasibility and evaluation of an emergency department-based GP streaming and treatment service

**DOI:** 10.1101/2022.05.13.22275043

**Authors:** Clare Aldus, Ian Pope, Julii Brainard, Annmarie Ruston, Gareth Hughes, Paul Everden

**Affiliations:** School of Health Sciences, University of East Anglia Norwich NR4 7TJ, UK; Norwich Medical School, University of East Anglia Norwich NR4 7TJ, UK; Emergency Department, Norfolk and Norwich University Hospital, Norwich NR4 7UY, UK; Faculty of Arts, Humanities, Canterbury Christ Church University, Canterbury Kent CT2 7NP, UK; North Norfolk Primary Care, Alkmaar House, Alkmaar Way, Norwich NR6 6BF, UK

**Keywords:** emergency department attendances, inappropriate attendances, service evaluation, treatment times, patient satisfaction

## Abstract

**BACKGROUND:** Emergency departments (EDs) are under ever-increasing pressure. The General Practitioner Streaming and Treatment (GPST) service implemented at a large ED in England UK aimed to identify and treat patients who attended an ED but who might effectively be managed in primary care to reduce pressure on ED services.

**METHODS:** Patients attending ED were met by a GP nurse practitioner who ‘streamed’ them to the GPST service or usual ED care. Routinely collected electronic records, satisfaction questionnaires and interviews were used to evaluate patient outcomes, staff experiences, service outcomes and impacts on usual ED services.

**RESULTS:** Approximately 96% of GPST patients were seen by a clinician within one hour and all within 87 minutes. Routinely collected ED datasets indicate statistically significant reductions in patients streamed to usual ED care who had to wait > 4 hours for disposition (p=<0.005). Of 769 patients with GPST consultation (approximately 10% of all walk-in patients) 421 (55%) needed no further intervention by ED. The speed at which GPST patients were managed exceeded patients’ expectations and was a major determinant of their satisfaction. No staff expressed dissatisfaction, but some suggested possible improvements in patient eligibility criteria and built environment design features.

**CONCLUSIONS:** Concurrent provision of GPST correlated with shorter waits for ED attenders to receive health care. Patient and staff experiences of GPST were positive. A robust assessment of safety and health economic outcomes would be useful to refine eligibility criteria and cost effectiveness.

## Introduction

Emergency Department (ED) services are under increasing pressure. In 2018-19 there were 24.8 million attendances in EDs in England. ^1^ This is an increase of 4 per cent compared with 2017-18 and 21 per cent since 2009-10.^1^ A 4-hour standard was introduced in the UK in 2004 for the largest EDs (“type 1”). It requires that 95 per cent of patients arriving at an ED should be admitted to hospital, transferred to a more appropriate care setting, or discharged home within four hours. Type 1 EDs in England have not met the 4-hour target at national levels since July 2013.^2^

The problem of breaching disposition targets is particularly acute at the Norwich and Norfolk University Hospital (NNUH) ED in eastern England, UK. The NNUH supports the oldest population in England, including North Norfolk district which has the oldest median age, at 53.8 years, of any local authority area in England and Wales. ^3^ The median resident age across Norfolk is about 45 years compared to median of 40.2 years for the UK. ^3^ Together, increased attendance and older median patient age have meant that the NNUH ED has particularly failed to meet the 4-hour target ^4^.

It has previously been identified that up to 54.2%,^5^ of patients attending EDs in high income countries may be suitable for primary care. This is not a new phenomenon. Cohen (1987) cited six studies that estimated between 35% and 89% of ED attendees were suitable for primary care management. ^6^ Streaming patients who could be managed elsewhere, away from or out of highly pressurised EDs to co-located GP-led primary care services, may support patients to receive the care they need whilst relieving pressure on ED services and improving ED performance against the 4-hour standard. Making it easy and easier for patients to obtain primary health care may be important to achieve resilience and adequate capacity in emergency health services ^7-10^. The UK National Health Service (NHS) currently offers a range of rapid access services to support EDs including walk-in centres, urgent care centres, minor injury units and urgent treatment centres. ^11^

A ‘feasibility study’ is a preliminary study. ^12^ Feasibility studies typically look at the design and acceptability of an intervention (service) and feasibility of wider testing or roll out and inform the design of further evaluative or definitive studies. This report describes a general practitioner

Streaming and Treatment Service (GPST) feasibility study conducted at the NNUH ED in the period 16 December 2019 to 28 February 2020. GPST aimed to improve patient experience; support access to appropriate care and resources; reduce the number of walk-in patients’ seen in ED; improve staff wellbeing; provide a safe service and change unplanned care to planned care where possible.

## Methods

### Design of the GPST service

The GPST was located at the front door of NNUH ED and aimed to improve patient experience, support access to appropriate care and resources, reduce the number of walk-in patients seen in ED, improve staff wellbeing, provide a safe service and change unplanned care to planned care where possible. It was delivered by general practice provider organisations: North Norfolk Primary Care, OneNorwich Practices and South Norfolk Healthcare having been commissioned by Norwich Clinical Commissioning Group (CCG), North Norfolk CCG and South Norfolk CCG.

The GPST process is illustrated in Figure 1. Briefly, trained clinicians screened and streamed patients who arrived at the ED using non-urgent modes of transport (ie. not conveyed via ambulance) as soon as possible after their arrival (ideally within 15 minutes). Streaming typically involved assessing eligibility via a brief history to determine whether the patient met the prescribed inclusion or exclusion criteria. Patients meeting inclusion criteria were offered the opportunity to book a same day appointment and follow concurrent NHS GP Improved Access Service Protocols ^13^.

**Figure 1.**
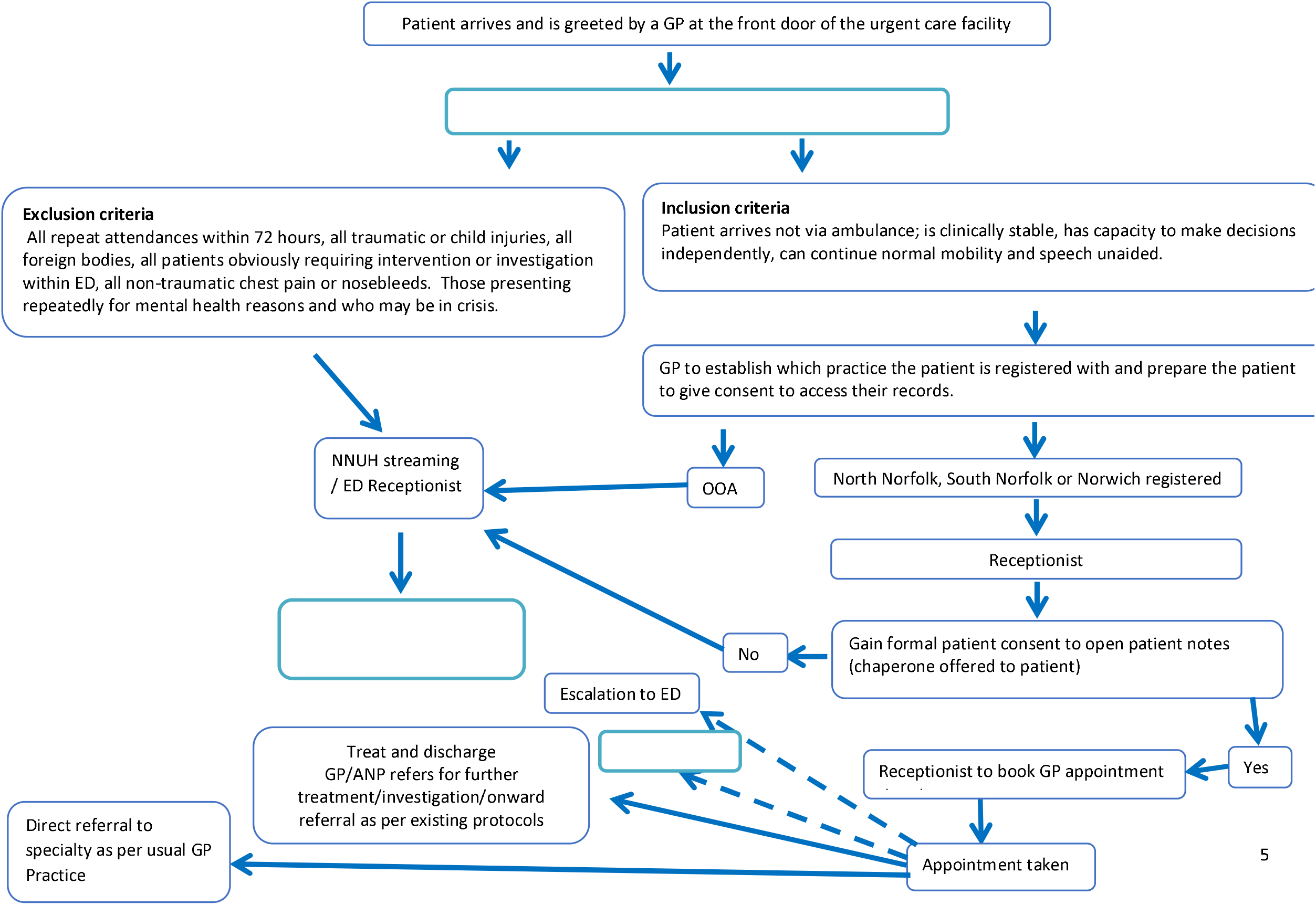
Patient pathway

Patients who were eligible clinically could continue to a GP appointment if they consented to GPST accessing their medical records. If an eligible patient deteriorated, they would be moved, via an agreed escalation route, to ED. Patients not eligible for inclusion in the GPST feasibility study were directed to usual ED streaming and care. The service was designed to operate between 8.30 am and 6 pm. Eligibility criteria were based on patient history, presentation and whether the attendee was registered at eligible GP surgeries (see Figure 1 for details).

## Aims of this service evaluation

To describe and evaluate the impact of the feasibility GPST service with respect to patients, staff and services. Evaluation objectives are:

1. To describe the activity undertaken by GPST
2. To assess whether patients are satisfied with their experience of GPST
3. To assess whether the presence of the GPST service has an impact on the wellbeing of staff who work there
4. To assess the potential reduction in burden on ED
5. To assess unintended consequences of GPST, such as increased use of other services or other adverse outcomes
6. To assess the ability of the service to change care from unplanned to planned
7. To assess whether the service is safe

## Evaluation methods

GPST feasibility study design was based on a Context, Mechanism, and Outcomes (CMO) framework to understand under which circumstances (context), how (mechanism), and for whom (outcomes) the service worked. ^14-17^ A range of measures were used to describe the activity, acceptability and impact of GPST. Evaluation was based on the GP STREAMING – TREATMENT evaluation plan Version 1.1 (Appendix 1), and conducted to determine the feasibility of GPST and to inform future evaluation. The GPST service and feasibility study commenced on 16th December 2019 running across 10 weeks to 28 February 2020.

### GPST activity

The number of patients seen by GPST; number of GP appointments available per week; number of advanced nurse practitioner appointments available per week; and the proportion of GPST appointments filled, not attended, or cancelled; were determined from routinely captured primary care electronic medical records (SystmOne ^18^) and staffing records.

### Patient and staff perceptions of GPST

Qualitative interviews were conducted amongst a sample of staff and patients. The study protocol (Appendix 1) proposed a draft topic guide. Fully authorised versions of topic guides used for patients and staff are presented in Appendix 2.

#### Patient interviews

At GP consultation a sample of patients were asked whether they would agree to be contacted about their experience of the GPST service. Patients who agreed to be contacted were invited to take part in a semi-structured telephone interview led by a researcher. Informed consent was obtained prior to the interview. Interviews were semi-structured and based on a topic guide (see authorised patient topic guide Appendix 2). The interviews were conducted at least one week after attendance at GPST by a research team member. Analyses were thematic and undertaken by the same researcher.

Patient participants were asked briefly about why they chose to use the GPST service; whether the service met their needs and expectations; and their experience. They were also asked about health outcomes (e.g. did you get better as a result?); and health service outcomes (did you need to visit another service as a result?); and whether they would use the service again; or recommend it to friends; and recommendations to improve the service.

#### Staff interviews

Informed consent was obtained prior to the interview. Interviews were semi-structured and based on a topic guide (see authorised staff topic guides Appendix 2). Semi-structured staff interviews were conducted by a member of the GPST team. Notes were recorded during the interview. Staff participants were asked about their experience of streaming patients; the prescribed eligibility criteria; consultations with patients; and repatriation of a patient to their usual GP. They were also asked about their perceptions of the purpose of GPST; and both positive and negative experiences and how the service or their experience may be improved.

### Patient satisfaction with GPST

Patients were asked to complete a brief questionnaire about the service. It comprised 4 items: patient experience, what could be improved, what do you plan to do next (about the reason for attendance), and any other comments.

### Staff wellbeing

Staff satisfaction and wellbeing was based on the validated Short Warwick-Edinburgh Mental Wellbeing Scale (SWEMWBS).^19 20^ The 7-item SWEMWBS presents a more restricted (largely psychological) view of mental wellbeing than the 14-item WEMWBS scale on which it is based but is more robust. ^20^ Dimensions included feeling optimistic about the future, feeling useful, feeling relaxed, dealing with problems well, thinking clearly, feeling close to other people, and being able to make up your own mind. Each item is scored from 1-5 therefore sum scores can range from 7 to 35.

### GPST impact

The impact of GPST was assessed in terms of the time to being seen by a doctor and number of breaches. Data used were routinely captured under electronic records held by primary care (SystmOne) or held by NNUH ED (Symphony). Data were used to assess whether the presence of GPST reduced burden on ED based on the number of walk-in patients seen in NNUH ED per day, time to being seen by a doctor and the number of breaches. Descriptive statistics and Student’s t-test (independent samples, two-tailed assuming unequal variance) at a 95% level of significance were used to describe and compare breaches in the 3 months prior to GPST deployment and during GPST implementation, including breakdown by patient category as ‘minors’, ‘majors’, ‘paediatric’ or ‘resus’.

### Unintended consequences

An unintended possible service impact was mean changes in attendance at a nearby (4 miles away) city centre walk-in health care clinic (Rouen Road), considering attendances both before introduction of GPST and concurrently on days that GPST was running. Changes in attendance for other services were not evaluated under this study. SystmOne ^18^, GPST and North Norfolk, South Norfolk and Norwich CCG data were used to determine the number of patients seen by a relatively nearby alternative walk-in centre (3.5 miles away, Rouen Road) who were subsequently referred to ED; and the number of patients referred from the co-located GP surgery for in-patient admission. Because it was the patients’ intention to access ED services these outcomes were not considered true unintended consequences and are reported under ‘GPST-impact’.

### Ability to change care from unplanned to planned

The number of patients who entered the GPST (planned) care pathway (i.e. were given a GPST consultation appointment) was determined as a proportion of the estimated number of NNUH ED walk-in patients for the evaluation period. The number of patients offered GPST appointments was determined from routinely captured SystmOne (primary care) electronic medical records and the estimated number of NNUH ED walk-in patients from Symphony (secondary care) records.

### GPST safety

Five patients who agreed to be interviewed were asked at telephone interview about unplanned representation to healthcare services within 48 hours and if repatriated back to their home practice for investigations whether they requested a GP appointment, or the required investigation was undertaken. GPST primary care staff were asked to feedback if they came across any safety issues in relation to use of GPST. The protocol was that if any eligible patient deteriorated whilst waiting for their GPST consultation appointment, they would be moved, via an agreed escalation route, to ED.

## Results

### GPST activity

GPST was designed to run from 08:30 to 18:00 hours on Monday to Friday from 16 December 2019 to 28 February 2020 with 40 appointment slots per day. It actually ran on 46 weekday dates across the 11-week period from 16 December 2019 to 28 February 2020 inclusive, excluding nine dates when insufficient staff were available: 19 and 23-27 December, 1-2 and 27 January.

Staff typically comprised a clinical GP, a streaming GP and an advanced nurse practitioner (ANP) across an 08:30 to 18:00 hr shift. Across the 46 dates, 39 (85%) dates were covered by a clinician GP, 43 (93%) by a clinician ANP; and 32 (70%) by a streaming GP. GPs and ANPs worked from 08:30 to 13:15 or 14:15 on a small number of service dates.

Typical patterns of attendance for patients seen in GPST is shown in Figure 2 by hour of attendance for all patients who were given a GPST consultation appointment. There appears to be reduced attendance in the middle of the day which corresponds closely with typical ED attendance patterns.

**Figure 2.**
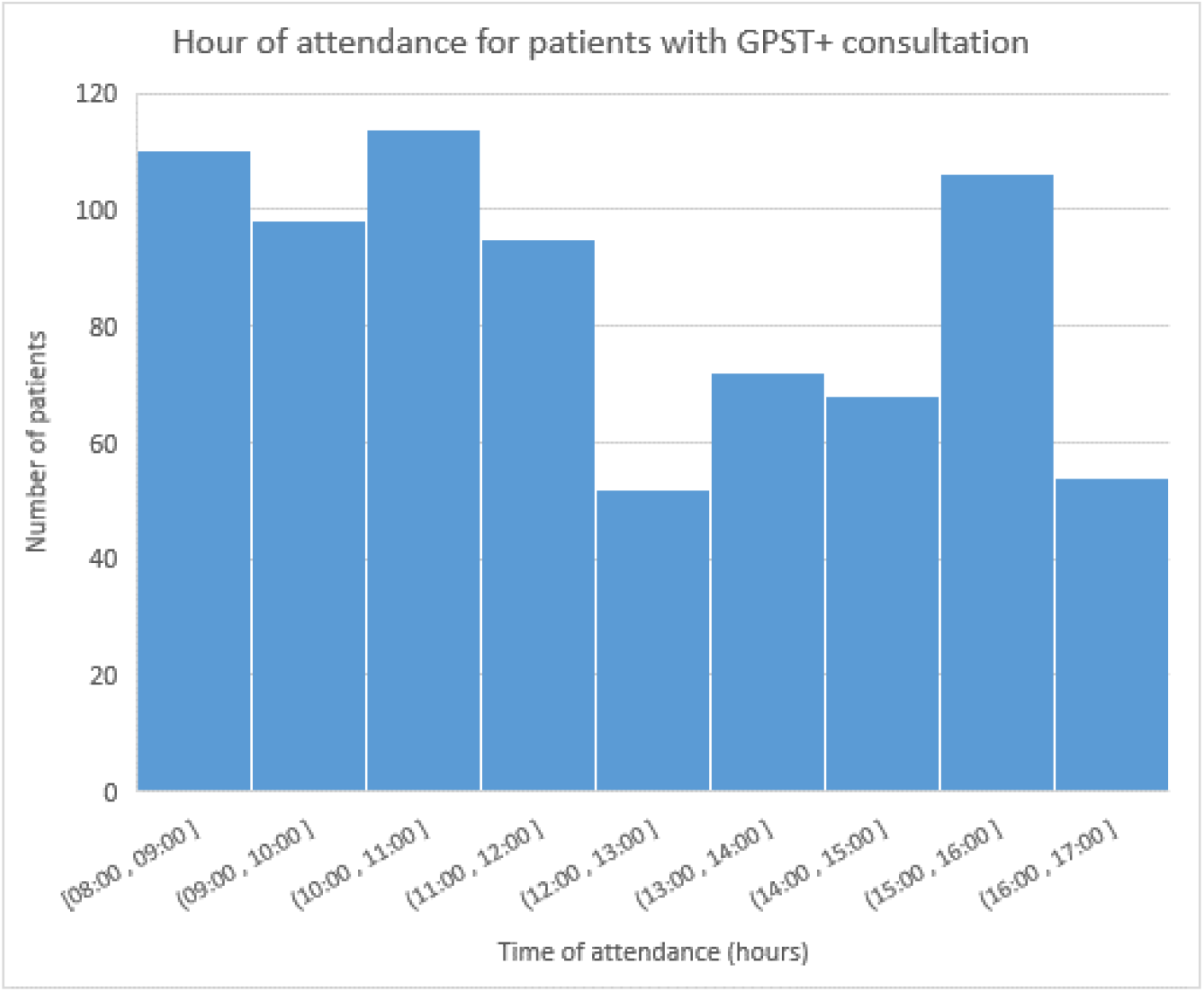
Distribution of attendance by hour for patients with GPST consultation

Data on the availability and utilisation of GP appointment slots were collected for 46 dates from 16 Dec 2019 to 28 Feb 2020 excluding weekends and 9 weekdays (19-December, 23-27 December (Christmas), 1-2 January (New Year), 27 January). The total number of appointments available was 3584 comprising 2415 GP appointments and 1169 ANP appointments.

Overall 768 (21.4%) of 3584 allocated slots were booked and 752 (21.0%) used. For GPs 449 of 2415 (18.6%) of allocated slots were booked and for ANPs the proportion was 319 of 1169 (27.2%). A chi-square test of independence with Yate’s correction ^21^ was performed to examine the relationship between profession (GP or ANP) and appointments booked or not booked. The relationship was significant (χ2=34.62, p=<0.001). ANP appointments were significantly more likely to be booked than GP appointments (p=0.05).

Patients generally attended their appointment slots. However, a small proportion of booked appointments were either not attended or cancelled. Overall 5 patients did not attend and 11 patients cancelled. The overall proportion not attended or cancelled was 16 (2.1%) of 768 booked appointments with 12 (2.7%) of 449 booked GP appointments not attended or cancelled and 4 (1.3%) of 319 for ANPs. Fisher’s exact test was used to examine the relationship between profession and appointments attended or cancelled/not attended. The relationship was not significant at p<0.05. ANP appointments were not more or less likely to be cancelled or unattended.

Across all patients booked for GPST appointments (n=769) mean waiting times were 13.9 (range 0 to 87) minutes (SD 15.65; 95% CI 12.82-15.04). The proportion of patients seen at their appointed time was not determined under this evaluation but of 769 patients given GPST appointments 325 (42.2%) were seen within 5 minutes, 449 (58.4%) within 10 minutes, and 666 (87%) within 30 minutes. Approximately 98% of patients were seen within one hour and all within 88 minutes. Seventeen patients (2.2%) waited for more than one hour to be seen. The distribution of elapsed times between patients arriving at ED (GPST screening) and their GPST consultation was provided and is illustrated in Figure 3 as a histogram of waiting times in 5-minute time bands and Figure 4 sequentially by arrival date and time.

**Figure 3.**
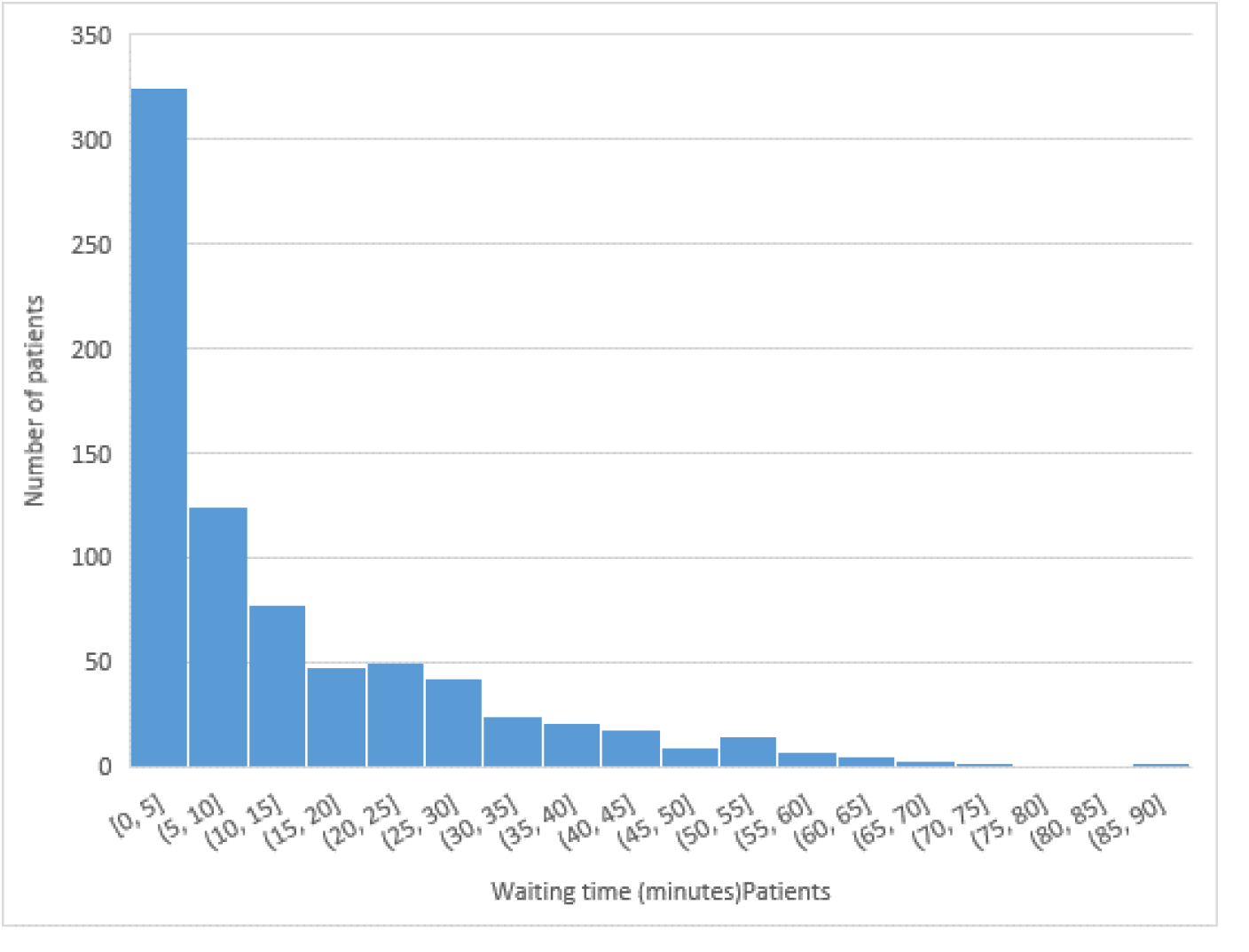
Distribution of waiting times for GPST consultation

**Figure 4.**
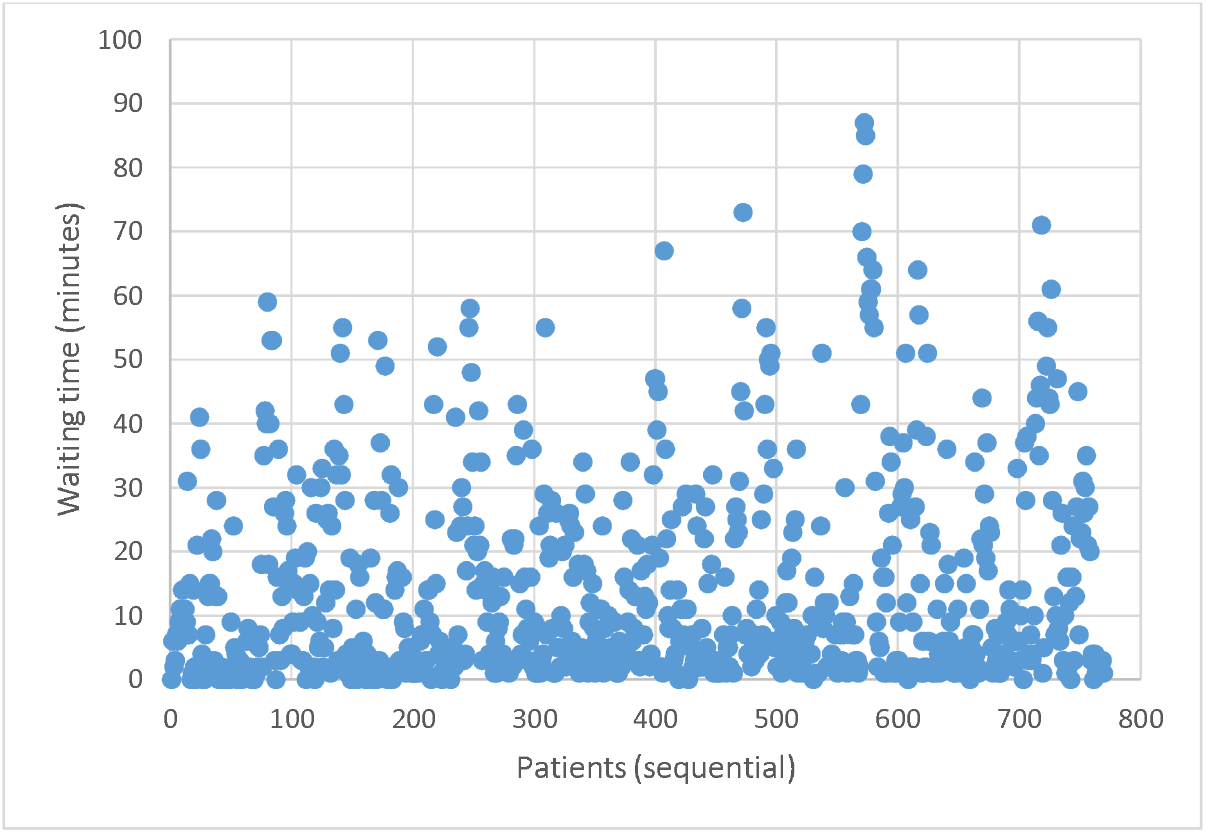
Elapsed times between patients arriving at ED (GPST screening) and GPST consultation sequential by date and time.

**Figure 5.**
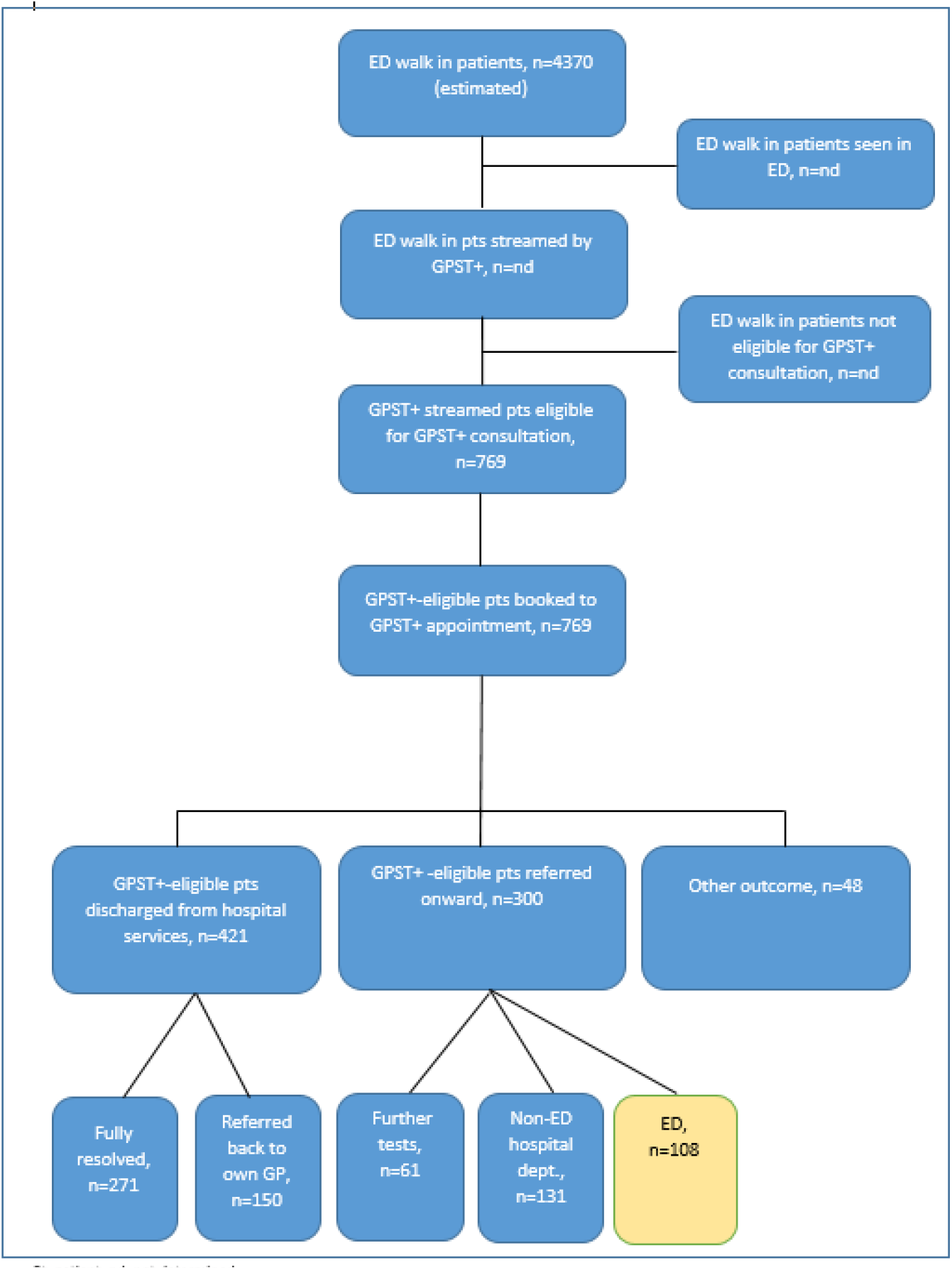
Flow of walk in patients attending NNUH ED during the GPST service evaluation

#### Patient and staff perceptions of GPST

##### Patient interviews

Twelve GPST adult patients and one adult acting as proxy for a child agreed to be contacted. Eight patients subsequently could not be contacted (n=3); did not respond to attempts to contact them (n=4); or did not agree to take part (n=1). Up to four attempts were made to contact each person. Five patients were interviewed. Interviews were conducted between 23 March 2020 and 30 March 2020 (at the start of COVID-19 lockdown).

Of those interviewed two said that they were unaware that they had agreed to be seen by GPST rather than usual ED care. The remaining three chose to use the service as an alternative to ED. Overall, respondents were positive about the service and three main themes emerged.

Firstly, the speed with which they were seen (compared with expectations):

> *“I visited the hospital because I had been to the GP when I thought I had pneumonia and with all the coughing my ribs were very painful. I wanted an Xray and expected to wait a long time but it only took five minutes. It was much better than I expected, I thought it would be four hours so really it exceeded my expectation*.*” Female Resp #5*
>
> *“I went to a pharmacist who said I had a dry eye and to see my GP but as it takes so long to see my doctor I decided to go to the hospital. It was great I was in and out in an hour and a half*.*” Male Resp #4*
>
> *“It was brilliant – it was better than waiting”. Female Resp #2*

Secondly, the perceived conditions that respondents felt the service was good for: conditions which ranged from straightforward to serious. This had the potential to influence their perspective of the service. For example, the following respondent had hurt her thumb when she fell over. She had been told by her GP to go straight to A&E:

> *“It is a good idea, this triage service and good for minor cuts, abrasions, minor falls – not major issues. It is good to separate out the cut thumb from something more serious. So it good for that…” Female Resp #3*.

Thirdly, that respondents may not necessarily know that they are using the service. For example, two respondents did not know that they had in fact been seen by the GP Streaming Service:

> *“I have a history of clots in my legs and chest and have had two pulmonary embolisms before. My chest was very tight and painful so I went to A&E but to be honest I did not know I had chosen that [GPST] service*.*” Female Resp #3*
>
> *“I don’t remember going in for that service but it was brilliant it was very quick. I had been to hospital two weeks before with a chest infection and because I have asthma I had to go back because it had not cleared up. I saw a nurse and she said she couldn’t hear anything in my chest but prescribed antibiotics*.*” Female Resp #1*

Overall, whether respondents were seen with potentially serious or minor conditions they appeared to be happy with the service they had received. No negative comments were received from service users.

#### Staff interviews

Six GPST clinicians were interviewed on 22 February 2020.

##### Experience of streaming

Clinician experience of streaming was generally positive and streaming was described as easy, interesting and working well. One clinician noted that GPST streaming provided a formal welcome to ED from a clinician and another that it enabled a clinician to meet immediate clinical need.

##### Experience of seeing patients

Clinicians described their experience of seeing patients as positive and similar to normal GP practice but one clinician identified that the environment limited confidentiality and sometimes therefore discussion could be uncomfortable. Clinicians felt that they were able to build rapport with the patient.

##### Views on exclusions

Inclusion and exclusion criteria are presented at Figure 1. GPST staff identified that the exclusion criteria were not always clear. For example ‘unstable’, ‘trauma’ or ‘mental health in crisis’ can present at a range of severity. Frustration with exclusion criteria was mentioned by one GPST member as a negative experience of GPST for them.

It was suggested that criteria surrounding trauma were too restrictive and that many more trauma cases such as wounds could have been seen. It was proposed that clinical judgement be allowed in assessing whether trauma cases could be managed by GPs and that staff be encouraged to manage trauma that they are comfortable with bearing in mind that skill sets vary. Also, it was unclear how dental problems should be managed.

One GPST staff member identified that children e.g. ‘foreign body in ear’ or ‘minor head injury’ could be safely managed by GPST with the ability to refer to the children’s ED. Patients who were eligible clinically could continue to a GP appointment if they consented to GPST accessing their medical records. One clinician noted a frustrating limitation surrounding electronic medical records e.g. the South Norfolk electronic medical records system (EMIS) led to exclusions which might otherwise have been managed by GPST.

##### Good use of resources

The cost of components of GPST was not presented so cost-effectiveness could not be evaluated but there was an overall view that GPST generally presented good use of resource. However, people identified that this could be improved if eligibility criteria were refined. Comments from elsewhere in interviews included use of GPST resources and NHS resources more broadly.

GPST staff were surprised at how many of the patients were sent to ED by GPs, many of whom had not been given a letter by their GP. And, where there was a GP letter, GPST staff were frequently unable to understand from the letter what the GP expected ED to do for the patient.

##### Delivering good clinical care

All respondents felt strongly that they could deliver good clinical care.

##### Experience of repatriation to GP

Some patients were referred back to their own GP. Five people responded with one person having no experience of this activity. All respondents were positive about their experience of repatriation to GPs with one commenting that the task was easy due to the systems in place.

##### Perceived purpose of GPST

GPST staff perceived the purpose of GPST was to reduce waiting time for patients (n=6), reduce pressure on ED (n=5), improve patient experience (n=4), improve patient care (n=2), save cost (n=2), or provide a safe service (n=1).

##### Experience of referral back to ED

Some GPST patients were referred back to ED care pathways. Four people commented on their experience of referral back to ED. Nobody had problems (n=4) “they [ED] just take them” and two indicated they felt well-supported in doing so. One person mentioned frustration that a patient would have to start their care pathway again.

##### Positive experiences

GPST staff communicated a range of positive experiences across two main themes. The first was about team relationships where they talked about the good communications and relationship between GPST and ED and the great experience of working together including breaking down barriers and misconceptions. The second was about professional satisfaction and patient care. Several mentioned that patients were happy with the care they had received. In accordance with the findings from patient interviews it was identified that patients were happy that, contrary to expectation, they were seen quickly. The fact that GPST enabled more streamlined care or made GPST staff feel they were getting things done was also raised. For example, one clinician relayed that for one sick patient they had used the internal phone to speak to an ED consultant. The consultant reserved a bed for the patient who went straight there.

##### Negative experiences

Negative experiences encompassed referrals, the physical environment, and clarity of roles. Several people mentioned frustration with the refusal of surgical units to accept referrals for patients. One person mentioned frustration caused specifically by a lack of response from eye casualty to telephone calls.

Specific aspects of the physical environment mentioned under negative experiences included [lack of] sinks, water temperature, lack of privacy at streaming. Also mentioned were water and room temperature. One person mentioned confusion of roles at streaming and suggested that unclear roles could cause friction.

##### Improvements

Suggestions for improvement were broadly in keeping with previous comments. They suggested improved privacy, better facilities (such as working sinks) and equipment e.g. a paediatric oxygen saturation monitor. They also suggested improvements to eligibility criteria and development of clear pathways. One person suggested increased staffing and opening hours to shift to slightly later in the day and another that patients and GPs more broadly should be educated on the purpose of ED.

#### Patient satisfaction with GPST

Of 769 patients booked for GPST appointment only four patient satisfaction questionnaires were returned. All four patients indicated high satisfaction. There were no relevant comments on improvements to the GPST service.

#### Staff wellbeing

The short Warwick-Edinburgh mental wellbeing scale (SWEMWS) was completed at least once by eight GPST staff with a mean score of 28.8 (range 28-35) for the first completion indicating good mental wellbeing for GPST staff. Two members of staff completed the SWEMWS on two and three separate occasions with mean scores of 34 and 35 respectively.

#### GPST impact

Outcomes for patients who were booked to attend GPST consultation (n=769) are described in Table 1. Outcomes can broadly be dichotomised into ‘managed in primary care’ (discharged from hospital services) or ‘managed in secondary care’ (referred onward to secondary care services) as presented in Figure 2. The 421 (55%) patients managed in primary care were either discharged ‘fully resolved’ (n=271, 64.4%) or referred back to their own GP (n=150, 35.6%). The 300 patients managed in secondary care were either referred for further tests (n=61, 20.3), or to non-ED hospital departments (n=131, 43.7%), or ED (n=108, 36%).

**Table 1.** Outcomes for GPST eligible patients as recorded by GPST

| Patient outcome | n (%) |
| --- | --- |
| Referred to ED | 108 (14.0) |
| No follow-up required | 271 (41.0) |
| Referred to own GP | 150 (22.7) |
| Referred for further investigation | 61 (9.2) |
| Referred to hospital | 131 (19.8) |
| Other outcome | 45 (6.8) |
| Other | 3 (0.1) |
| **Total** | **769 (100)** |

It is unclear from these analyses whether some of the ‘further tests’ were also undertaken in primary care. Outcomes for 45 (5.8%) patients were categorised as ‘other’ outcome and include 16 (2%) patients who did not attend or cancelled their appointment. Four patients (included in the referred to hospital services group) were admitted to hospital services: Early Pregnancy Assessment Unit (n=1); Acute Medical Unit (n=2); Gynaecology (n=1).

Figure 6 describes daytime walk-in ED attendances for the years 2018=2020. The average number of walk-in attendances between 8 am and 4 pm on weekdays in January and February 2020 was 95 in 2020 compared to 102 in 2019 and 85 in 2018. On average 17 patients (769 patients across 46 dates) were given GPST appointments each day. Based on an average of 95 walk-in patients attending NNUH each day it is estimated that almost one fifth (18%) of daily walk-in patients attended GPST appointments. Of these more than half (55%) were managed entirely within primary care services. Comparing groups there was no evidence of any impact of GPST on use of walk-in centres (p=0.150) with on average 159.1 (range 115-215) and 164.8 (range 83-226) daily attendance pre-GPST and during GPST respectively.

**Figure 6.**
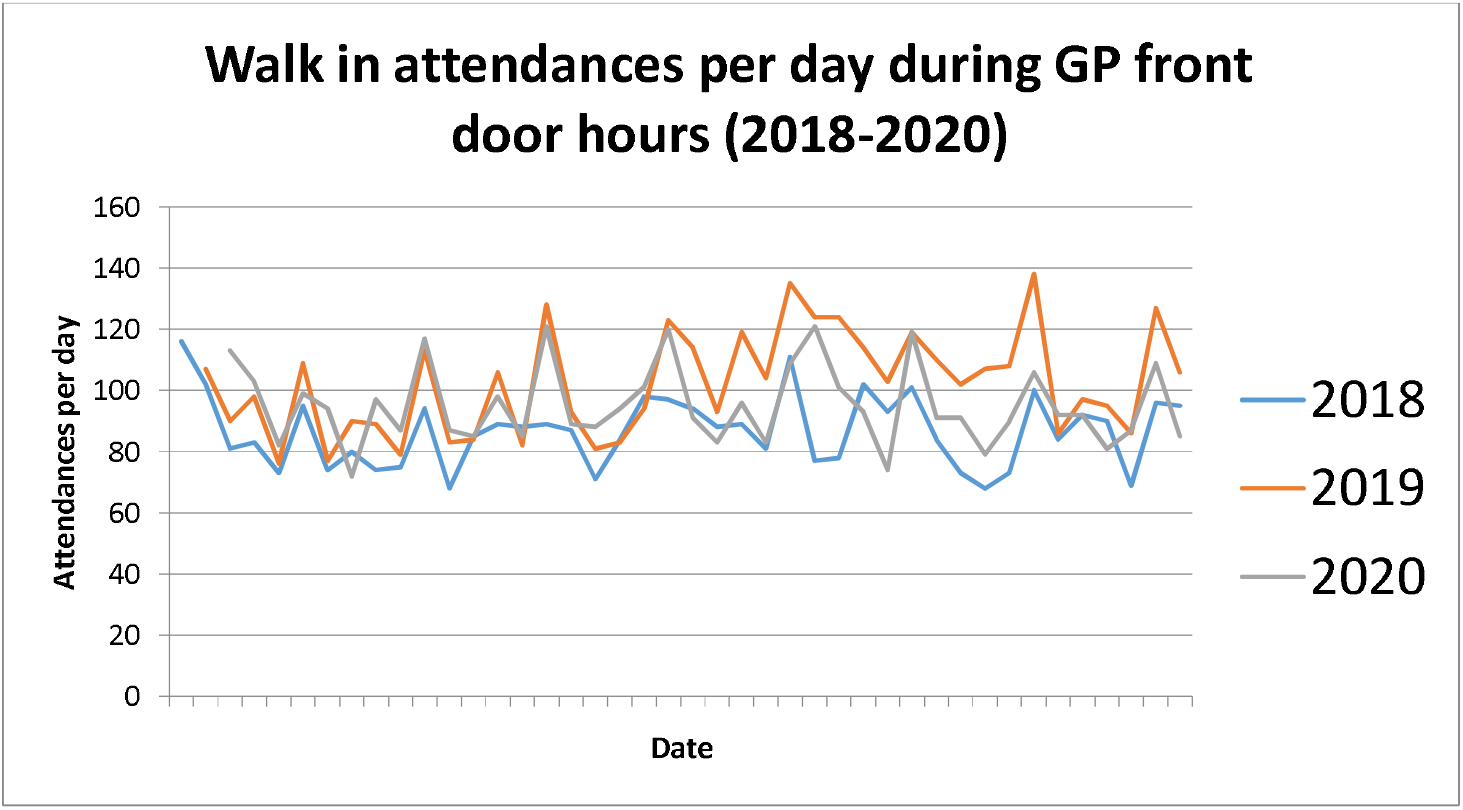
NNUH ED Walk in attendances 0800 to 1600 hours week days 01 January to 28 February 2020

Routinely collected ED data indicate that ‘round the clock’ for three months prior to implementation of GPST (01 Sept – 13 Dec 2019 excluding weekends) 29.4% of all patients were seen by a doctor in <60 minutes compared to 37.8% during-GPST (16 December 2019 – 28 February 2020 excluding weekends).

Figure 7 presents findings for weekday breaches by ED triage category pre- and during implementation of GPST. It is important to note that the results are for 24-hour breaches but GPST only worked during the day (approx. 9.00 to 17.00 h). Furthermore, GPST was not operational at weekends (not shown) or on nine weekdays (shown: 19-December, 23-27 December (Christmas), 1-2 January (New Year), 27 January). It indicates a close correlation between a reduction in absolute number of breaches and implementation of the GPST service (broken arrow).

**Figure 7.**
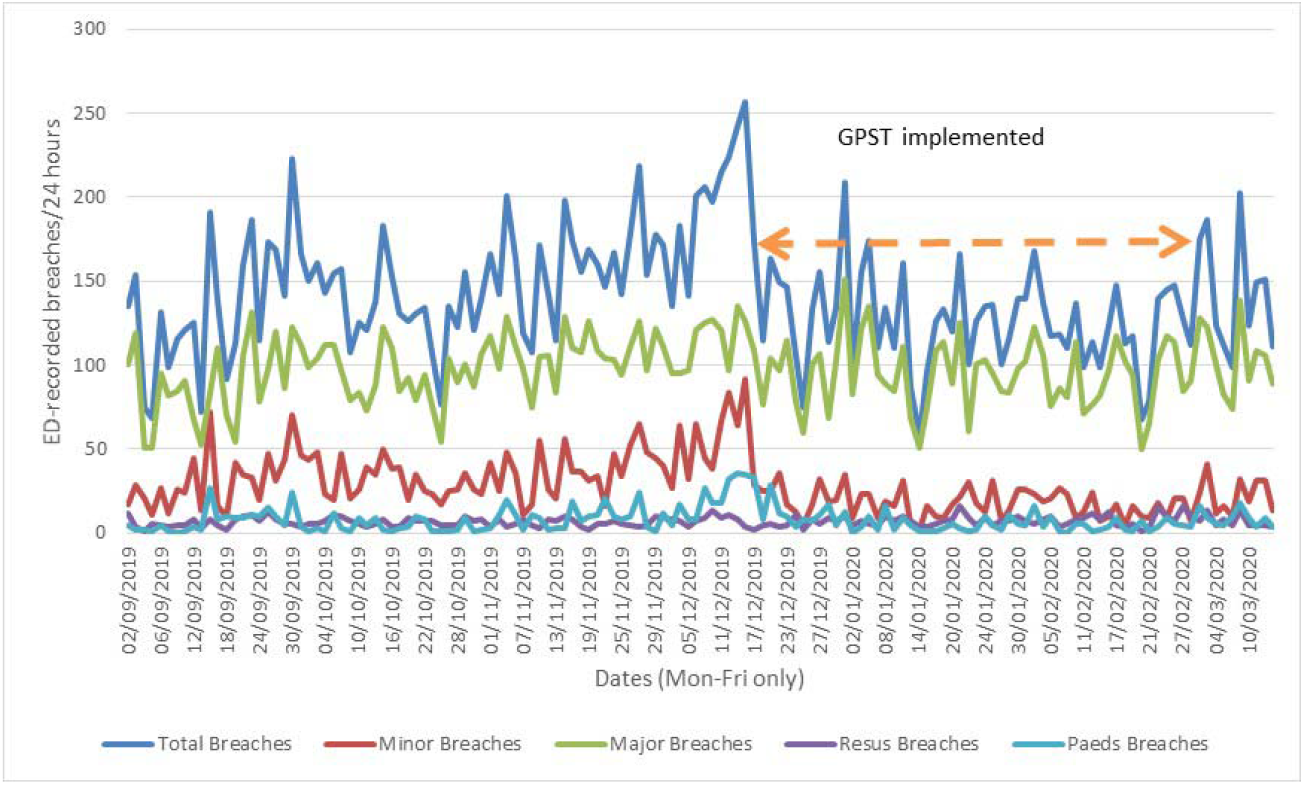
Round the clock weekday breaches by ED triage category pre-, during- and post-implementation of GPST (GPST implementation period indicated by arrow)

Table 2 describes the mean number and percentage change in number of breaches by triage category for the three-month pre-GPST period (September 1 to December 13 2019) and the GPST implementation period (December 16 to February 28). It indicates a reduction in the mean number of breaches overall (16.3%); for patients categorised as ‘minor’ (51.2%) paediatric (27%) and a small reduction for ‘majors’ (5%).

**Table 2.**
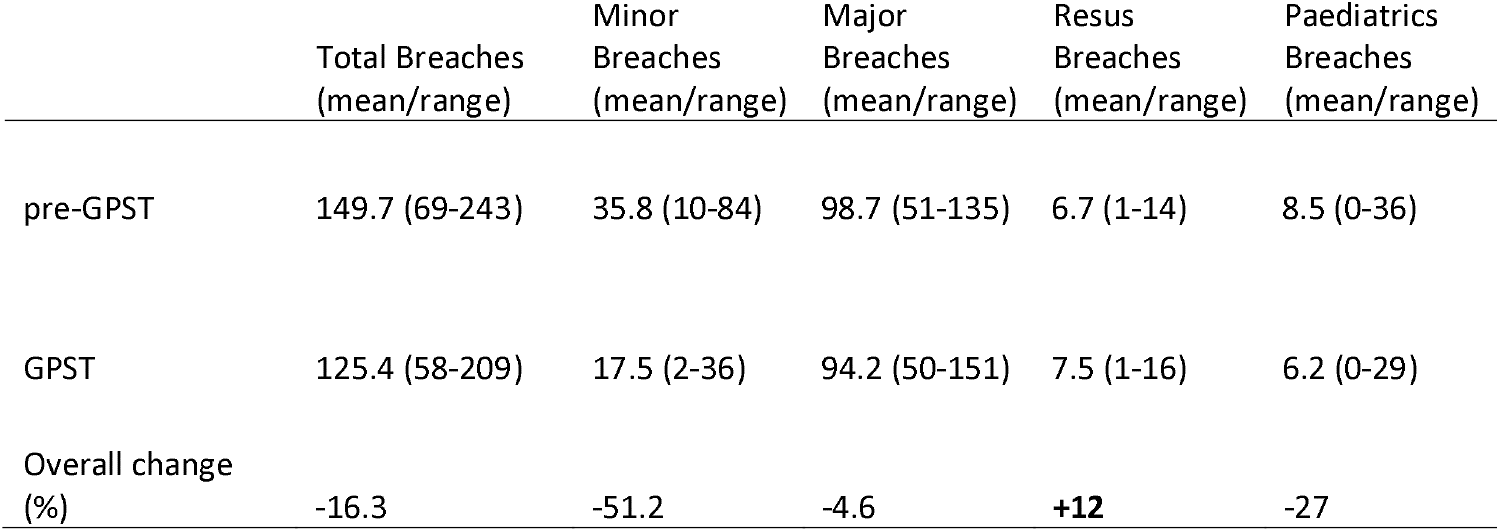
Mean number and percentage change in number of breaches by triage category for the period prior to and during GPST.

|  | Total Breaches<br>(mean/range) | Minor<br>Breaches<br>(mean/range) | Major<br>Breaches<br>(mean/range) | Resus<br>Breaches<br>(mean/range) | Paediatrics<br>Breaches<br>(mean/range) |
| --- | --- | --- | --- | --- | --- |
| pre-GPST | 149.7 (69-243) | 35.8 (10-84) | 98.7 (51-135) | 6.7 (1-14) | 8.5 (0-36) |
| GPST | 125.4 (58-209) | 17.5 (2-36) | 94.2 (50-151) | 7.5 (1-16) | 6.2 (0-29) |
| Overall change<br>(%) | -16.3 | -51.2 | -4.6 | **+12** | -27 |

Comparing ED breaches pre- and concurrent with GPST indicates statistically significant reduction in the mean number of breaches overall (p=0.003), highly significant reduction for minor breaches (p=<<0.005) and no significant change in breaches between groups for ‘majors’ (p=0.519); ‘resus’ (p=0.252) or ‘paeds’ (p=0.430) for the concurrent GPST group compared to the pre-GPST group.

#### Ability to change care from unplanned to planned

Of an estimated 4370 NNUH ED weekday daytime walk in patients (average of 95 patients per day across 46 dates) 769 (17.6%) agreed to attend GPST: i.e. they changed from unplanned to planned care. However, the total number of patients streamed by GPST (including those who did not enter GPST) was not captured under this evaluation. It is important to note that during busy periods it is possible that not every daytime walk-in patient was streamed by GPST. Therefore, the rate of conversion from unplanned to planned care as a result of the GPST streaming intervention remains unclear.

#### GPST safety

No GPST staff reported any patient safety issues in relation to use of GPST. However, lack of handwashing facilities, presenting a potential safety issue, were reported. No patients were escalated from GPST to ED pathways, due to patient deterioration, as potentially anticipated in the protocol.

## Discussion

GPST ran throughout the planned period except on 19 December and 27 January due to an inability to staff the service. Around 85% of dates were covered from 8.30 am to 6 pm with the remainder covered from 8.30 am to early afternoon (1.15 pm or 2.15 pm). The ability to staff the service should be a consideration in future service design.

One important aspect of feasibility for NHS services is to determine flow through the service. 3584 GPST appointment slots were potentially available for an estimated 4370 ED walk in patients i.e. capacity to provide 82% of all NNUH walk in patients with GPST appointments. Overall approximately 21% of the potential appointment slots were allocated to around 18% of the daytime ED walk-in patients. It is important to note that estimates from previous studies clearly suggest that more than 18% of patients are appropriate for GP-led care ^5, 6, 22^. The findings from this study may suggest that there was an over-supply of appointment slots or that there was insufficient capacity to manage more patients due to e.g. eligibility criteria, patient condition, flow, space or equipment or that only 18% of patients attending were suitable for GPST. In reality the answer probably lies somewhere in between. Management of resource and revised eligibility criteria should be carefully considered prior to further evaluation. This could be supported by an evaluation of reasons for attendance and which reasons are suitable for the GPST service. Process evaluation should be conducted to determine how effectively GPST and ED protocols are delivered.

GPST was effective in seeing 42%, 58% and 87% of patients within 5, 10 and 30 minutes respectively. Approximately 96% of patients were seen by a doctor within one hour and all within 87 minutes. Separate routinely collected ‘round the clock’ ED data suggests that on average there was an 8% increase in patients seeing a doctor within 60 minutes during the GPST period indicating improved service outcomes.

A breach occurs where a patient is not admitted to hospital, transferred to a more appropriate care setting, or discharged home within four hours of arrival. GPST collected data indicates that 421 (55%) of 769 patients (approximately 10% of all walk-in patients) needed no further intervention by ED and that no GPST patients breached the 4-hour target. Thus it seems reasonable to assume that the GPST service which managed around one fifth (18%) of walk-in patients may potentially translate into reduced breaches in ED and may potentially improve the capacity of ED to manage the remainder of ED patients.

This study was not a comparative evaluation but did explore the number of breaches by triage category before and during GPST implementation. Breach data routinely collected by NNUH ED indicates a close correlation between a reduction in overall breaches and ‘minors’ breaches for the dates that GPST was implemented. The close correlation in service design and dates implemented strongly suggest but do not confirm causation. However, the fact that changes in the mean numbers of breaches for ‘majors’, ‘resus’ and ‘paeds’ (none of which were the target of the GPST intervention) were not statistically significant strengthens the case for causation. In addition, breach data provided and reported herein, were for ‘round the clock’ attendances and if assessed for daytime GPST hours only, would likely further underpin the likelihood of causation.

Future evaluation should robustly explore how GPST impacts the capacity of NNUH ED to meet its 4-hour targets through comparative evaluation which includes cost-effectiveness analysis. There was no evidence of any negative impact of GPST on use of walk-in centres. Acceptability is an important aspect of any feasibility study. Patients indicated that the GPST service was acceptable to them and staff also expressed satisfaction with their experience of working in the service. There was no evidence to suggest that patients or staff did not find the GPST service acceptable and the short Warwick-Edinburgh mental wellbeing scale (SWEMWS) completed by eight GPST staff with a mean score of 28.8 (range 28-35) indicated good mental wellbeing for GPST staff.

Most patients (98%) attended their booked appointments thus providing quantitative data indicating that GPST was acceptable. Qualitative data indicated that the speed at which patients were managed exceeded their expectations (also identified in staff interviews) and seemed to be a major determinant of their satisfaction. They could also see that GPST provided a service appropriate to minor complaints. However, some patients were also unaware that they had attended the GPST service. This may indicate rapid assimilation of GPST within ED but equally may indicate that patients were unfamiliar with usual ED practice. However, it should be noted that very few patients (n=5) were interviewed or returned satisfaction surveys (n=4).

GPST staff wellbeing and professional satisfaction was good as indicated by wellbeing surveys and staff interviews. However, the built environment was criticised by GPST staff. The main problem was a lack of working sinks but minor equipment and the temperature of the rooms was also mentioned. Related to the built environment a key concern was a lack of privacy during patient screening where sensitive concerns and ‘controlling’ carers were noted.

The design of further evaluation should include robust and systematic collection of patient and staff satisfaction data. The design should consider alternative methods to deliver questionnaires or recruit subjects for interview. Interviews with patients and staff receiving GPST or ED care should be recorded, analysed thematically and transcribed. Ethnography surrounding streaming and care pathways for ED and GPST would be valuable.

The impact of GPST was primarily determined through exploration of patient pathways. Around 55% of patients (n=421) allocated to GPST appointments were entirely managed within primary care. This is around 10% of the total number of walk-in patients for the study period. A further 39% (n=300) patients were referred for other tests or to secondary care services including ED. The remainder were categorised as ‘other outcomes’. Only 14% of patients allocated to GPST consultation appointment reverted to ED services. Thus 86% of patients meeting all GPST eligibility criteria and 15% of the estimated number of patients overall (661 of 4370) were removed from the ED pathway.

The number of patients streamed to GPST appointment and subsequently referred to ED (108 of 769) seems relatively high. Ethnography and analysis of the condition of patients that fall into this group may add value to any future study and reduce the burden for patients who, if referred to ED, need to restart their patient journey again.

Under this evaluation not all aspects of patient flow were captured. The design of a new study should capture all stages of patient flow e.g. how many patients were excluded form GPST at each of the eligibility stages. In a future evaluation to support full details of all stages of the onward journey must be effectively captured to reduce the size of any unclear ‘other outcomes’ category and to confirm e.g. whether further tests are carried out in primary or secondary care. This is particularly important in determination of comparative effectiveness and cost effectiveness analyses.

No patients were escalated from GPST to ED pathways as a result of deterioration in health (as per protocol) during their GPST consultation appointment. This may suggest that eligibility criteria are effective with respect to safety. However, along with comments from staff, and the relatively low proportion of patients (18%) streamed to GPST appointments (compared to evidence from research studies) this finding may indicate that eligibility criteria are over cautious. GPST staff thought that eligibility criteria should be loosened and allow more freedom for clinical judgement.

This evaluation of the GPST service was designed to focus on GPST. However, feasibility studies should also inform future study design. The design of a future evaluation should include robust comparative evaluation of ‘ED with GPST’ and ‘ED without GPST’ to underpin evidence-based commissioning decisions. A range of designs are possible but this may be achieved by a before-and-after study or by random allocation of a proportion of walk-in ED attendees to one or other service. Quantitative and qualitative research should be designed to include both services.

### Strengths and limitations

The GPST service and feasibility study was co-designed by a team from primary care, secondary care and healthcare commissioning services with **a priori** development of this feasibility study. Patient decisions to attend A&E and staff availability may have been affected because the last few weeks of the service coincided with the start of the UK Covid-19 epidemic. The numbers of patients who returned questionnaires or were interviewed about their experience of GPST was low so these results should be considered tentative. Cost-effectiveness was not undertaken.

## Conclusion

The findings from this feasibility study suggest that: GPST can successfully identify patients suitable for streaming to primary care; manage patients in a timely fashion; successfully reduce ED burden; and provide a safe service. GPST provided an acceptable alternative to ED care for patients who used it and staff who worked in the service. Further evaluation is not ruled out on the grounds of safety or acceptability for staff and patients and therefore a pilot study should be designed to enable robust comparative evaluation of ‘ED with GPST’ and ‘ED without GPST’ including effectiveness and cost effectiveness.

We recommend that the design of the built environment must be fit for GP-at-door services. Prescribed inclusion and exclusion criteria should be carefully selected. The degree of clinical judgement surrounding eligibility, which is acceptable and beneficial to GPST and ED services could be explored further. Clinical pathways which help to clarify and streamline GPST and ED roles and services should be developed.

## Data Availability

All data are patient specific, sensitive and not available publicly.

## Acknowledgements

We thank the patients and staff of GPST and NNUH ED for their support.

## Appendix 1. Protocol for GPST service EVALUATION PLAN

### Purpose

This protocol outlines proposals for an independent evaluation led by the University of East Anglia (Faculty of Medicine and Health Sciences) of an initiative locating the GP Streaming – Treatment+ service in front of the Emergency Department (ED) at Norfolk and Norwich University Hospital (NNUH). This is in the context of increased waiting times and the perception that some attendances could be better managed by services other than ED. Hospitals across England are increasingly unable to meet the “four hour target”, which assesses departments on the proportion of patients arriving and leaving the ED in less than 240 minutes. Streaming patients who could be managed elsewhere, away from or out of highly pressured EDs, to co-located GP led primary care services, may support patients receiving the care they need whilst improving performance against the four-hour standard.

The service is commissioned by Norwich CCG, North Norfolk CCG and South Norfolk CCG and was initiated to support improved access to appointments in the community and reduce burden on ED. The service will initially be a 6-8-week pilot, which commenced on 17^th^ December 2019. The purpose of this evaluation is inform future decisions about investment and service design.

### Description of proposed pilot service

#### Purpose of service

Objectives of the service:

- Improve patient experience;
- Support access to appropriate care and resources;
- Reduce the number of walk-in patients’ seen in ED;
- Improve staff wellbeing;
- Provide a safe service
- Change unplanned care to planned care where possible.

### The service

Trained clinicians will screen patients at the front door of ED as soon as possible after their arrival (and always within 15 minutes). Streaming will typically involve a brief history and use of inclusion/exclusion criteria to assess if a patient is suitable for a GP appointment. If they are suitable, they will be offered the opportunity to book a same day appointment and follow the GP Improved Access Service Protocols. If the patient deteriorates, they will move to an agreed escalation route to ED. Patients not suitable for the GP STREAMING - TREATMENT+ route will be directed to the ED Streaming Nurse/Reception and continue the ED patient pathway in the normal way. This process is summarised in Figure 1 below. The service operates between 8.30am and 5.30pm.

**Figure 1.**
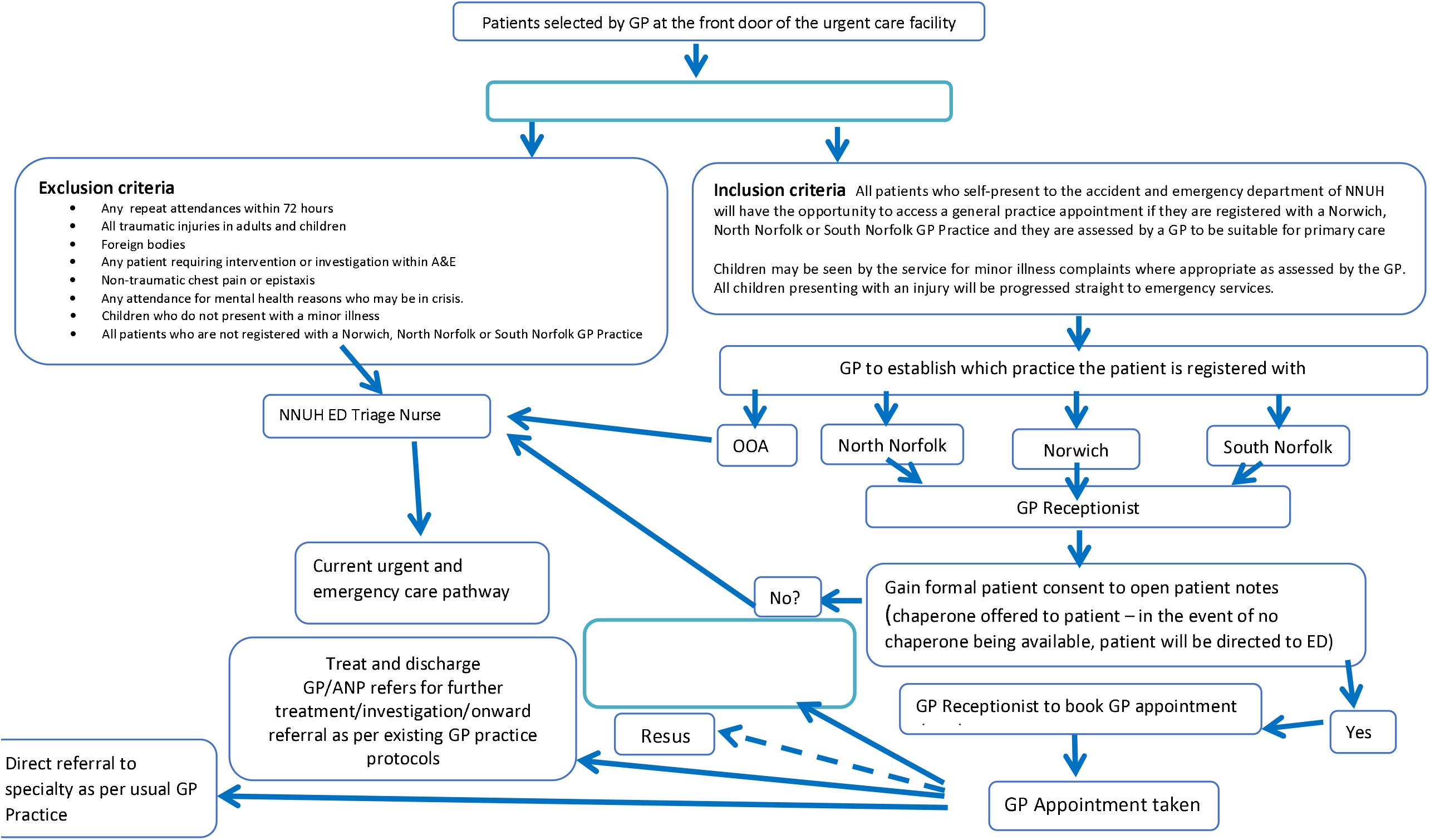
GPST eligibility and pathways. Notes: OOA, Out of area (patients not registered with GP surgeries in North Norfolk, Norwich or South Norfolk); Resus, resuscitation stream of ED; Majors, major trauma stream of ED

### Aims and Objectives of this Evaluation

#### Aim

To evaluate the impact of the GP Streaming – Treatment+ on patient care, patient satisfaction, staff wellbeing and NNUH ED.

#### Objectives

8. To describe the activity undertaken by GP Streaming – Treatment+.
9. To assess whether patients are satisfied with their use of GP Streaming – Treatment+.
10. To assess whether the presence of the GP Streaming – Treatment+ service has an impact on staff wellbeing.
11. To assess whether the presence of GP Streaming – Treatment+ reduces burden on ED.
12. To assess unintended impacts of the service (e.g. increased use of other services, adverse events).
13. To assess the ability of the service to change care from unplanned to planned.
14. To assess whether the service is safe.

### Methods

This protocol was informed by the NHS/NIHR “Evaluation Works” service evaluation toolkit (Evaluation Works 2019) and “Evaluating Interventions” (Ovretveit 1998). As the period for the pilot evaluation is short (6-8 weeks) we will focus on the collection and analysis of routinely collected data. Where resources allow, we will also collect data to support the evaluation objectives.

### Evaluation framework

A realist evaluation framework will be used to make sense of quantitative and qualitative data (Pawson & Tilley 1997; Pawson 2006). Realist frameworks evaluate what works (outcome) for who in what circumstances (context) and how (mechanism). This is known as a ‘CMO’ (Context, Mechanism, Outcome) framework (Figure 2).

**Figure 1.**
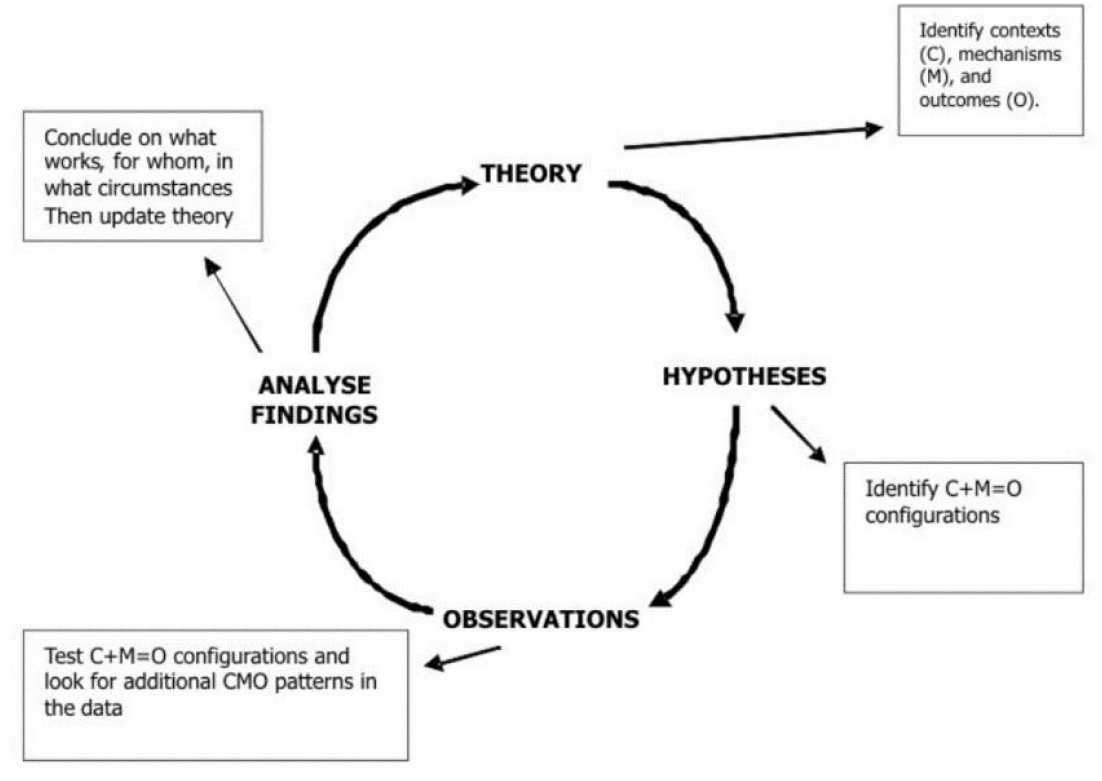
The realist evaluation cycle (Pawson & Tilley 1997)

### Quantitative measures

#### Routine data

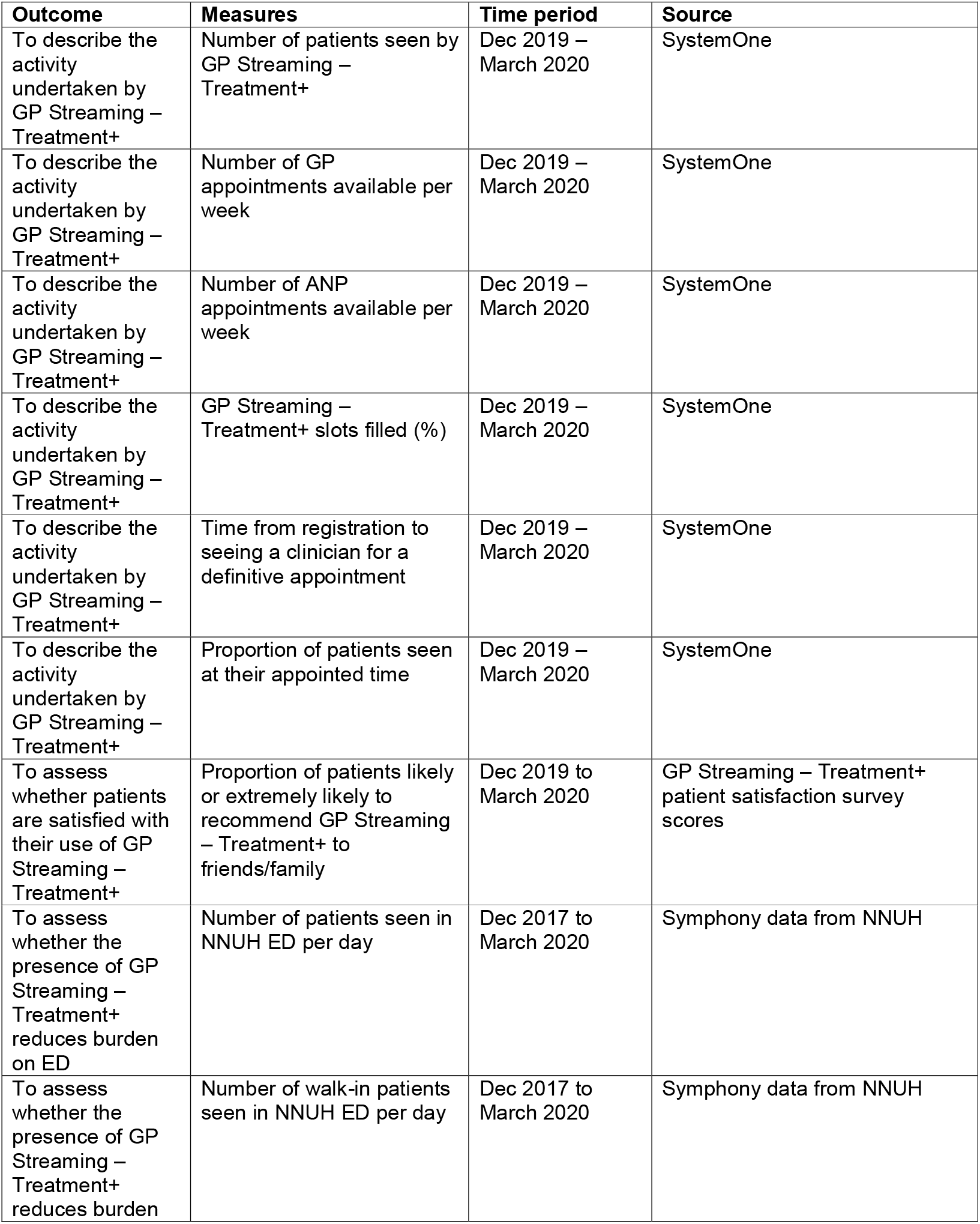
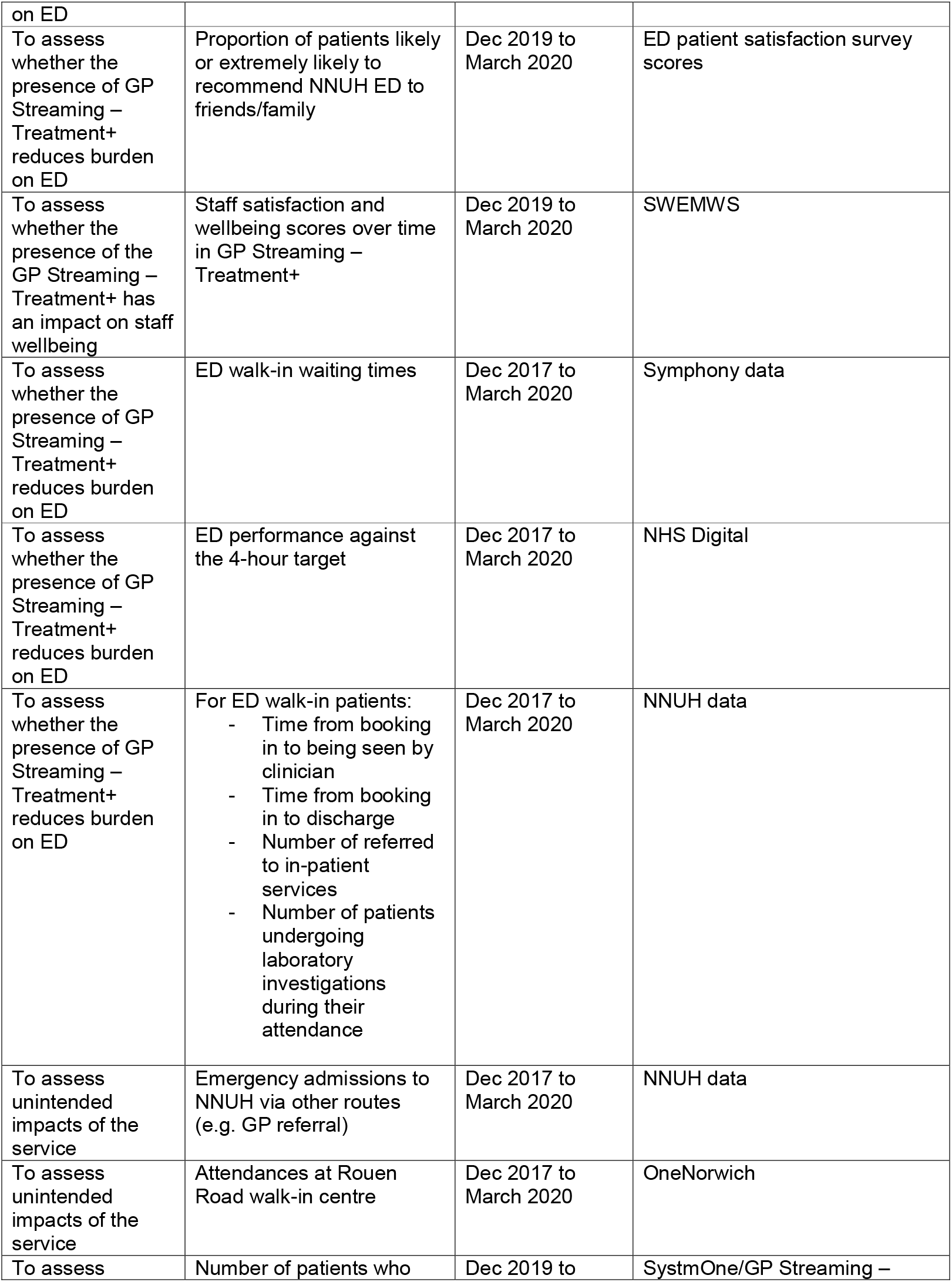
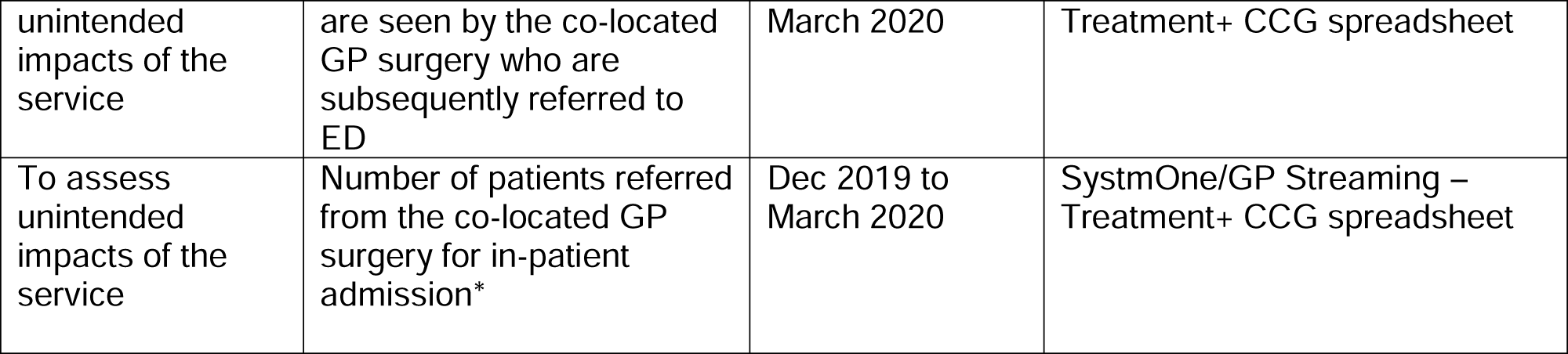

#### Other data

| Outcome | Measures | Time period | Source |
| --- | --- | --- | --- |
| Ability to change care from unplanned to planned. | Number of patients discharged from NBPM who have an unplanned re-presentation to healthcare services within 48 hours | Dec 2019 – March 2020 | Phone calls to a sample of patients |
| Ability to change care from unplanned to planned. | Number of patients repatriated back to their home practice for investigations or referral who subsequently have the requested investigation or referral made | Dec 2019 – March 2020 | Phone calls to a sample of patients |
| To assess whether the service is safe. | To collect data on adverse events we will ask practices to feedback if they come across any in relation to use of GP Streaming – Treatment+ | Dec 2019 – March 2020 | Primary care staff |

#### Qualitative data

Qualitative interviews will be conducted with a sample of staff and patients. Informed consent will be obtained prior to the interview.

Interviews will be semi-structured and based on a topic guide. We expect the topic guide to include:

- Does the process of GP streaming effectively identify suitable patients to be seen in GP Streaming – Treatment+? Are GPs the most suitable clinical workforce for the presenting conditions seen?
- Could the process of streaming to GP Streaming – Treatment+ be done more effectively?
- What information do GPs gather during the streaming process and does it change management?
- Which patients are most suitable for the GP Streaming – Treatment+?
- How do clinicians find the process of seeing ED patients in GP Streaming – Treatment+? How do clinicians find the process of repatriating patients back to their own GP practice?
- How do patients perceive being seen in GP Streaming – Treatment+?
- What are the barriers to streaming patients to GP Streaming – Treatment+?
- Are there more patients who are suitable to be seen in GP Streaming – Treatment+ that are currently being seen?
- Do the location and times of operation seem appropriate? Why/why not?
- Does the unit address a need that A&E at NNUH is not set up to manage?
- Barriers and facilitators to pathways and flow between departments
- Clinician perceptions as to which groups of patients can be seen safely in this service and which cannot

#### Analysis

Interviews and focus groups will be recorded where possible, or extensive notes taken. A thematic analysis will then be undertaken.

### Timelines

The pilot service went live on 17^th^ December 2019 and will run for 6-8 weeks. This evaluation will be conducted concurrently, and we aim to provide a short evaluation report at the end of March 2020.

## Appendix 2. Approved feasibility study documentation

### Appendix 2.1: Participant Information Sheet

Version 28/2/20

**Service Evaluation of GP Front Door/Streaming Treatment and Service Project**

#### Service Evaluation

##### Participant Information Sheet

We would like to invite you to take part in our service evaluation. This information sheet will provide you with details about the aims of the evaluation as well as what we will ask of you. Please take your time to read the following information. If you have any questions, or would like more information, please feel free to contact us.

###### What is the purpose of this evaluation?

The aim of this evaluation is to examine the effect of the GP Front Door/Streaming Treatment and Service on patient care and satisfaction, staff well being and the organisation and delivery of Emergency Department Services.

###### Why have I been invited?

You have been invited as you are either:

a. A patient that has used the service.
b. A member of staff who has been involved in the delivery of care or the organisation or management of the service.

###### Do I have to take part?

No, your participation is voluntary. You are free to withdraw from the interviews at any point. If you agree to an interview and you withdraw, your data will be destroyed up to the point at which it is anonymised

###### What does the project involve for me?

The study will involve either a face-to-face meeting within the Emergency Department or a telephone/face to face interview at a time and place that is convenient for you. The interview would last about thirty minutes.

###### What will you do with my information?

We will record the conversation either on a tape recorder or on paper. Your data will be anonymised and stored securely on university servers. Your consent form will be kept in a locked cabinet in a locked office at UEA in accordance with the university’s data protection rules.

###### Will my information be confidential?

Yes. Your involvement in the evaluation will remain strictly confidential and all data will be kept on a password protected computer and accessed by the research team only.

###### Complaints and further contact

If you have any further questions about this evaluation or any of the information given above please do not hesitate to contact us on the e-mail addresses below and we will be happy to answer any questions you may have. If you have any concerns or complaints about the research then please contact the Professor Dylan Edwards, Dean of the Faculty of Medicine and Health Sciences

###### Contact details

Principal investigators: Dr Ian Pope

Clinical Lecturer in Medical Education

Norwich Medical School,

University of East Anglia,

Norwich,

Telephone number: 01603 597191

Dr Paul Everden

Honorary Senior Lecturer

Norwich Medical School,

University of East Anglia,

Norwich,

### Appendix 2.2: Consent form

Version 1; 28/2/20

Service Evaluation of GP Front Door/Streaming Treatment and Service Project

#### Interview consent form

If you agree to participate in the above evaluation please **initial** in the box at the end of each sentence and complete the details at the bottom of the form.

|  |  |
| --- | --- |
| 1 | I confirm that I have read and understood the information sheet for participants, dated 16.12.19 version 1, and have had the opportunity to ask questions. |
| 2 | I agree to the interview being audio recorded and/or written notes taken for the purposes of analysis and possible publication. |
| 3 | I understand that all data will be anonymised and any identifiable factors will be removed by the evaluation team. |
| 4 | I agree to be interviewed to participate in this study. |
| 5 | I understand that I can withdraw my consent to participate at any point but that my data may not be removable. |

\_\_\_\_\_ Name of participant
\_\_\_\_\_ Signature
\_\_\_\_\_ Date
\_\_\_\_\_ Name of person taking consent

### Appendix 2.3: Indicative topic guide for Clinicians Screening Patients

Version 1; 28.2.20;

*Introduction*

*Overview of evaluation aims and objectives*

*Structure of interview and reconfirmation of consent*

Could you describe your role within the delivery/organisation of the service?

What do you think the aim/purpose of the service is?

Could you describe your experience of streaming patients into the service?

Has your experience been what you expected or having you found anything surprising?

What are your views on the inclusion and exclusion criteria?

Are there any specific conditions the service could see but are currently excluded?

If you were running the service what would you do differently?

What is your view on whether the service is a good use of resources?

Have you enjoyed working in the service – is there anything that could be done to make your experience of working in the service better?

If you were asked to advise the organisers of the service on how the service could be improved what three things would you recommend?

Anything you would like to add?

Thank you very much for taking your time to participate. All data will be processed confidentially and we can send you a copy of the report if you would like to know the outcomes of the project.

### Appendix 2.4: Indicative topic guide for Clinicians Seeing/Treating Patients

Version 28; 8.2.20;

*Introduction*

*Overview of project aims and objectives*

*Structure of interview and reconfirmation of consent*

Could you describe your role within the delivery/organisation of the service?

What do you think the aim/purpose of the service is?

Could you describe your experience of /seeing treating patients using the service?

Has your experience been what you expected or having you found anything surprising?

What are your views on the inclusion and exclusion criteria?

Are there any specific conditions the service could see but are currently excluded?

Could you describe a particularly positive experience that you have had whilst working in the service?

Could you describe a particularly negative experience that you have had whilst working in the service?

What is your view on whether the service is a good use of resources?

How would you describe the quality of clinical care provided by the service?

What has your experience of repatriating patients back to their own GP investigations or follow up been like – could you give examples?

What has your experience of referring patients on to the ED been like – could you give examples?

If you were asked to advise the organisers of the service on how the service could be improved what three things would you recommend?

#### Closing question

Anything else you would like to add?

#### Ending

Thank you very much for taking your time to participate. All data will be processed confidentially.

### Appendix 2.5: Indicative topic guide for Receptionists booking patients into the service

Version 1; 28.2.20;

*Introduction*

*Overview of project aims and objectives*

*Structure of interview and reconfirmation of consent*

Could you describe your role within the delivery/organisation of the service?

What do you think the aim/purpose of the service is?

Could you describe your experience of booking patients into the service?

Has your experience been what you expected or having you found anything surprising?

Have you faced any particular difficulties in undertaking this role?

If you were asked to advise the organisers of the service on how the service could be improved what three things would you recommend

#### Closing question

Anything else you would like to add?

#### Ending

Thank you very much for taking your time to participate. All data will be processed confidentially and we can send you a copy of the report if you would like to know the outcomes of the project.

### Appendix 2.6: Indicative topic guide for patients immediately after seeing a clinician from the service

Version 1; 28.2.20;

*Introduction*

*Overview of project aims and objectives*

*Structure of interview and reconfirmation of consent*

What made you decide to choose to use the service today?

How was your experience of contact with the people on the front desk offering the service?

What was your experience of being seen by one of the clinicians?

Did it match your initial expectations? *if yes why? If no why?*

If you came in again with the same problem would you still choose to use the service, *if yes why? If no why?*

What do you think would have improved the service for you?

What do you plan to do next with regards to the problem you came in with today?

#### Closing question

Anything else you would like to add?

#### Ending

Thank you very much for taking your time to participate. All data will be processed confidentially.

### Appendix 2.7: Indicative topic guide for ED streaming nurses

Version 1; 28.2.20;

*Introduction*

*Overview of project aims and objectives*

*Structure of interview and reconfirmation of consent*

Could you describe your role within the delivery/organisation of the service?

What do you think the aim/purpose of the service is?

How have you found the GP front door service? Do you think they are seeing appropriate patients?

If you were asked to advise the organisers of the service on how the service could be improved what three things would you recommend?

### Appendix 2.8: Indicative topic guide for ED management

Version 1; 28.2.20;

*Introduction*

*Overview of project aims and objectives*

*Structure of interview and reconfirmation of consent*

Could you describe your role within the delivery/organisation of the service?

What do you think the aim/purpose of the service is?

How do you think it is performing currently?

What effect do you feel it is having on ED workload? How will we know if the service is performing well? How do you think it could be improved?

If you were asked to advice on how it could be improved what three things would you recommend?

#### Closing question

Anything else you would like to add?

#### Ending

Thank you very much for taking your time to participate. All data will be processed confidentially.

### Appendix 2.9: Indicative topic guide for GP streaming – treatment and management

Version 1; 28.2.20;

*Introduction*

*Overview of project aims and objectives*

*Structure of interview and reconfirmation of consent*

Could you describe your role within the delivery/organisation of the service?

What do you think the aim/purpose of the service is?

How do you think it is performing currently?

What effect do you feel it is having on ED workload?

How will we know if the service is performing well?

How do you think it could be improved?

If you were asked to advice on how it could be improved what three things would you recommend?

### Appendix 2.10: Indicative topic guide for patients after their attendance (minimum one week after)

Version 1; 28.2.20;

*Introduction*

*Overview of project aims and objectives*

*Structure of interview and reconfirmation of consent*

What made you decide to choose to use the GP Front Door service?

Did it meet your needs on the day? *If yes why and if no why*

How was your experience of contact with the people on the front desk offering the service?

What was your experience of being seen by one of the clinicians?

Did it match your initial expectations? *if yes why? If no why?*

Following your visit to the service what happened next?

- *Did it clear up*
- *Did you need to visit another service to follow up?*
- *Did you have to return to ED?*

If you needed follow up investigations or referrals did these happen and how did it work for you?

If you came in again with the same problem would you still choose to use the service, *if yes why? If no why?*

Would you recommend the service to others? *If yes why if no why?*

What sort of conditions do you think should be seen by the service?

If you were asked to advise the organisers of the service on how the service could be improved what three things would you recommend?

What do you think would have improved the service for you?

